# Country wide surveillance reveals prevalent artemisinin partial resistance mutations with evidence for multiple origins and expansion of high level sulfadoxine-pyrimethamine resistance mutations in northwest Tanzania

**DOI:** 10.1101/2023.11.07.23298207

**Authors:** Jonathan J. Juliano, David J. Giesbrecht, Alfred Simkin, Abebe A. Fola, Beatus M. Lyimo, Dativa Pereus, Catherine Bakari, Rashid A. Madebe, Misago D. Seth, Celine I. Mandara, Zachary R. Popkin-Hall, Ramadhan Moshi, Ruth B. Mbwambo, Karamoko Niaré, Bronwyn MacInnis, Filbert Francis, Daniel Mbwambo, Issa Garimo, Frank Chacky, Sijenunu Aaron, Abdallah Lusasi, Fabrizio Molteni, Ritha J. A. Njau, Samwel Lazaro, Ally Mohamed, Jeffrey A. Bailey, Deus S. Ishengoma

**Affiliations:** University of North Carolina, Chapel Hill, NC, USA; Brown University, Providence, RI, USA; National Institute for Medical Research, Dar es Salaam, Tanzania; Nelson Mandela African Institute of Science and Technology, Arusha, Tanzania; Harvard T.H Chan School of Public Health, Boston, MA, USA; Broad Institute, Boston, MA, USA; National Malaria Control Programme, Dodoma, Tanzania; Swiss Tropical Public Health Institute, Dar es Salaam, Tanzania; Muhimbili University of Health and Allied Sciences, Dar es Salaam, Tanzania; Faculty of Pharmaceutical Sciences, Monash University, Melbourne, Australia

## Abstract

**Background:** Emergence of artemisinin partial resistance (ART-R) in *Plasmodium falciparum* is a growing threat to the efficacy of artemisinin combination therapies (ACT) and the efforts for malaria elimination. The emergence of *Plasmodium falciparum* Kelch13 (K13) R561H in Rwanda raised concern about the impact in neighboring Tanzania. In addition, regional concern over resistance affecting sulfadoxine-pyrimethamine (SP), which is used for chemoprevention strategies, is high.

**Methods:** To enhance longitudinal monitoring, the Molecular Surveillance of Malaria in Tanzania (MSMT) project was launched in 2020 with the goal of assessing and mapping antimalarial resistance. Community and clinic samples were assessed for resistance polymorphisms using a molecular inversion probe platform.

**Findings:** Genotyping of 6,278 samples collected countrywide in 2021 revealed a focus of K13 561H mutants in northwestern Tanzania (Kagera) with prevalence of 7.7% (50/649). A small number of 561H mutants (about 1%) were found as far as 800 km away in Tabora, Manyara, and Njombe. Genomic analysis suggests some of these parasites are highly related to isolates collected in Rwanda in 2015, supporting regional spread of 561H. However, a novel haplotype was also observed, likely indicating a second origin in the region. Other validated resistance polymorphisms (622I and 675V) were also identified. A focus of high sulfadoxine-pyrimethamine drug resistance was also identified in Kagera with a prevalence of dihydrofolate reductase 164L of 15% (80/526).

**Interpretation:** These findings demonstrate the K13 561H mutation is entrenched in the region and that multiple origins of ART-R, similar as to what was seen in Southeast Asia, have occurred. Mutations associated with high levels of SP resistance are increasing. These results raise concerns about the long-term efficacy of artemisinin and chemoprevention antimalarials in the region.

**Funding:** This study was funded by the Bill and Melinda Gates Foundation and the National Institutes of Health.

**Research in Context:** *Evidence before this study:* We did a literature search via PubMed for research articles published from January 2014 to October 2023 using the search term “Africa” and “Artemisinin resistance” linked to “R561H” or “A675V” or “R622I”, returning 32 studies. The published literature shows the emergence and establishment of these three validated *Plasmodium falciparum* kelch13 (K13) mutations associated with artemisinin partial resistance (ART-R) in Africa. Large molecular studies of 675V in Uganda and 622I in Ethiopia have defined the regional spread of these mutations. However, limited data is available from recent studies about the spread and origins of the 561H mutation in the Great Lakes region of East Africa. In particular, detailed studies of the regions of Tanzania that border Rwanda have not been carried out since the mutation was detected in Rwanda. These data are needed for malaria control programs to define and implement strategies for controlling the spread of ART-R in Africa, a potential global public health disaster and the potential obstacle to the ongoing elimination strategies.

*Added value of this study:* This analysis reports the first large-scale analysis of antimalarial resistance in Tanzania, with a focus on the regions bordering Rwanda since the 561H mutation reached high frequency in the area. Using 6,278 *P. falciparum* positive samples sequenced using molecular inversion probes (MIPs), we show that the mutation has become frequent in the districts of Kagera bordering Rwanda. Importantly, we provide evidence for the separate emergence of a different extended haplotype around 561H in Tanzania. This is the first evidence that multiple independent emergences of the 561H ART-R have occurred in Africa, as was seen within the last two decades in Southeast Asia.

*Implications of all the available evidence:* These findings highlight that, similar to 622I and 675V in other parts of Africa, we can expect the 561H mutation to continue to spread in the region. In addition, it highlights that we need to be watchful for new origins of mutations beyond the spread of existing resistant parasite lineages. ART-R appears to now be well established in multiple areas in Eastern Africa. Intensive control in these regions to prevent spread and monitoring for partner drug resistance emergence in affected areas will be critical for preventing further reversal of malaria control efforts in the region and support progress to the elimination targets by 2023.

## INTRODUCTION

Resistance to drugs used for treatment and prevention of malaria poses one of the greatest threats to global control and is of grave concern in Africa where the vast majority of cases and deaths occur.^1^ Historically, the emergence and spread of chloroquine and sulfadoxine-pyrimethamine (SP) resistance resulted in the collapse of effective treatment of malaria with significant increases in morbidity and mortality.^2^ Antimalarial combination therapies, consisting of an artemisinin derivative and a partner drug (artemisinin-based combination therapy - ACTs), are currently the predominant therapeutic options for treating uncomplicated falciparum malaria. Given gains in malaria control have already plateaued and are reversing in some countries, the emergence of artemisinin partial resistance (ART-R) in Africa could be a global public health disaster if partner drug resistance emerges in concert resulting in frank ACT failure.^3^

Over a decade has passed since the emergence of ART-R in SouthEast Asian *P. falciparum* populations, leading to decreased drug efficacy.^4^ Clinical ART-R was first demonstrated in the late 2000s in studies conducted in Western Cambodia. The emergence of ART-R in Western Cambodia set the stage for the eventual failure of ACTs, as resistance to the partner drugs also emerged, indicated by increasing treatment failures, parasite clearance times and partner drug IC50s over a very short timeframe.^3^ Early in the emergence of ACT resistance, many areas of SEA experienced more than 50% failure in patients treated with ACTs due to the combined effect of artemisinin and partner drug resistance.^3^

Mutations in *Plasmodium falciparum* Kelch13 (K13) (PF3D7_1343700) are the key mediator of ART-R. These mutations were originally identified through drug pressure experiments and validated in the field and by genetic engineering studies.^5^ They are thought to alter ubiquitination patterns and help parasites to resist accumulation of polyubiquitinated proteins.^5^ A number of key mutations in K13 propeller domain are now validated markers of ART-R. Unfortunately, validated K13 mutations have now been found extensively in Eastern Africa, particularly in the Horn of Africa (R622I), Uganda (C469Y, A675V), Rwanda (R561H), and Tanzania (R561H).^6–10^

The regional emergence and spread of these mutations appears to be developing as the norm rather than the exception. In the Horn of Africa, the validated 622I mutation was first reported in 2014.^11^ Since that time, these mutations have spread across Eritrea and Ethiopia.^7,8^ In Uganda, longitudinal molecular surveillance at 16 sites has painted a clear picture of spread within Uganda of the 469Y and 675V variants.^6^ It is important to note that all three of these mutations have been associated with prolonged parasite clearance, day 3 positivity by microscopy, increasing prevalence over time, and are in regions that now meet WHO criteria for ART-R.^7,8^

The emergence and spread of the 561H mutation in the Great Lakes Region of East Africa is not yet as clear when compared to other mutations. Originally described in Rwanda in samples from 2014 and 2015, this mutation appears to have emerged within the country over the past decade.^10,12^ In 2015, 7.4% of samples collected in Masaka harbored the 561H mutation.^10^ By 2018, 561H prevalence had increased to 19.6% in Masaka and 22% in Rukara during a therapeutic efficacy study.^13^ In this study, 50% of isolates with delayed clearance (day 3 positive parasitemia) carried 561H. On this basis, Masaka met the WHO criteria for endemic ART-R as defined by >5% of patients carrying K13 resistance-confirmed mutation for which they had persistent parasitemia by microscopy on day 3.^14^ Genome sequencing of isolates from 2015 also confirmed a single haplotype of 561H in Rwanda that was not of Asian origin, suggesting *de novo* mutation within Africa.^10^

Importantly, K13 mutations should never be studied in isolation as it requires partner drug resistance to lead to clinical failures of ACTs. Further, mutations to drugs no longer used for therapy, but that remain in use for chemoprevention (e.g. SP), are also important to characterize for malaria control programs. Molecular surveillance of antimalarial resistance should therefore rely on platforms that can broadly detect different resistance mutations. Highly multiplex amplicon deep sequencing is one approach that has shown promise. Another approach is molecular inversion probes (MIPs), which have now been used extensively to characterize drug resistance and population structure in parasites in Africa.^8,9,15^ The ability to create and combine different highly multiplexed panels for antimalarial resistance mutations, copy number variation, gene deletions and other genome wide polymorphisms to study complexity of infection, parasite relatedness or population structure makes the MIP platform a highly flexible and cost-effective means of conducting malaria molecular surveillance (MMS).

The Molecular Surveillance of Malaria in Tanzania (MSMT) project was developed to provide nationwide longitudinal surveillance of parasite populations to understand key aspects of parasite biology that may impact malaria control and interventions.^16^ The emergence of 561H is a prime concern for Tanzania given its proximity to Rwanda and previous studies documenting isolated cases across the country, with two cases in the Chato district and one on the Eastern coast.^9,17^ Here we describe the initial assessment of 6,278 successfully genotyped samples collected across 7,782 malaria positive individuals in the first year of the project using high-throughput MIP analysis. The goal was to assess the status of antimalarial resistance in Tanzania, with a focus on the border with Rwanda, to understand the distribution of the 561H mutation, partner drug resistance and resistance to chemoprevention drugs. This was further supported by whole genome sequencing of specific isolates to understand flanking haplotypes and gain insight into the origins of 561H mutations in Tanzania.

## MATERIALS AND METHODS

### Study Design and Participants

We analyzed dried blood spot (DBS) samples from patients with a positive malaria rapid diagnostic test drawn from cross-sectional surveys that involved 100 health facilities in 10 regions (n=7,148) and asymptomatic individuals in community studies (n=634) in three additional regions (**Figure 1**). Details of the study sites’ selection and overall sampling have been provided elsewhere.^16^ The three community surveys were conducted in regions of Tanzania which were involved in previous studies undertaken by the National Institute for Medical Research (NIMR).^9,18^ Four regions (Kagera, Mara, Tabora, and Kigoma) were deemed high priority areas for antimalarial resistance surveillance due to their proximity to Rwanda and to their location in the Lake Zone of the country with higher transmission (**Figure 1**). Informed consent was obtained for each patient and de-identified DBS samples were processed at NIMR in Tanzania, Brown University, and University of North Carolina (USA) according to IRB requirements of the Tanzanian Medical Research Coordinating Committee (MRCC) of NIMR.

**Figure 1.**
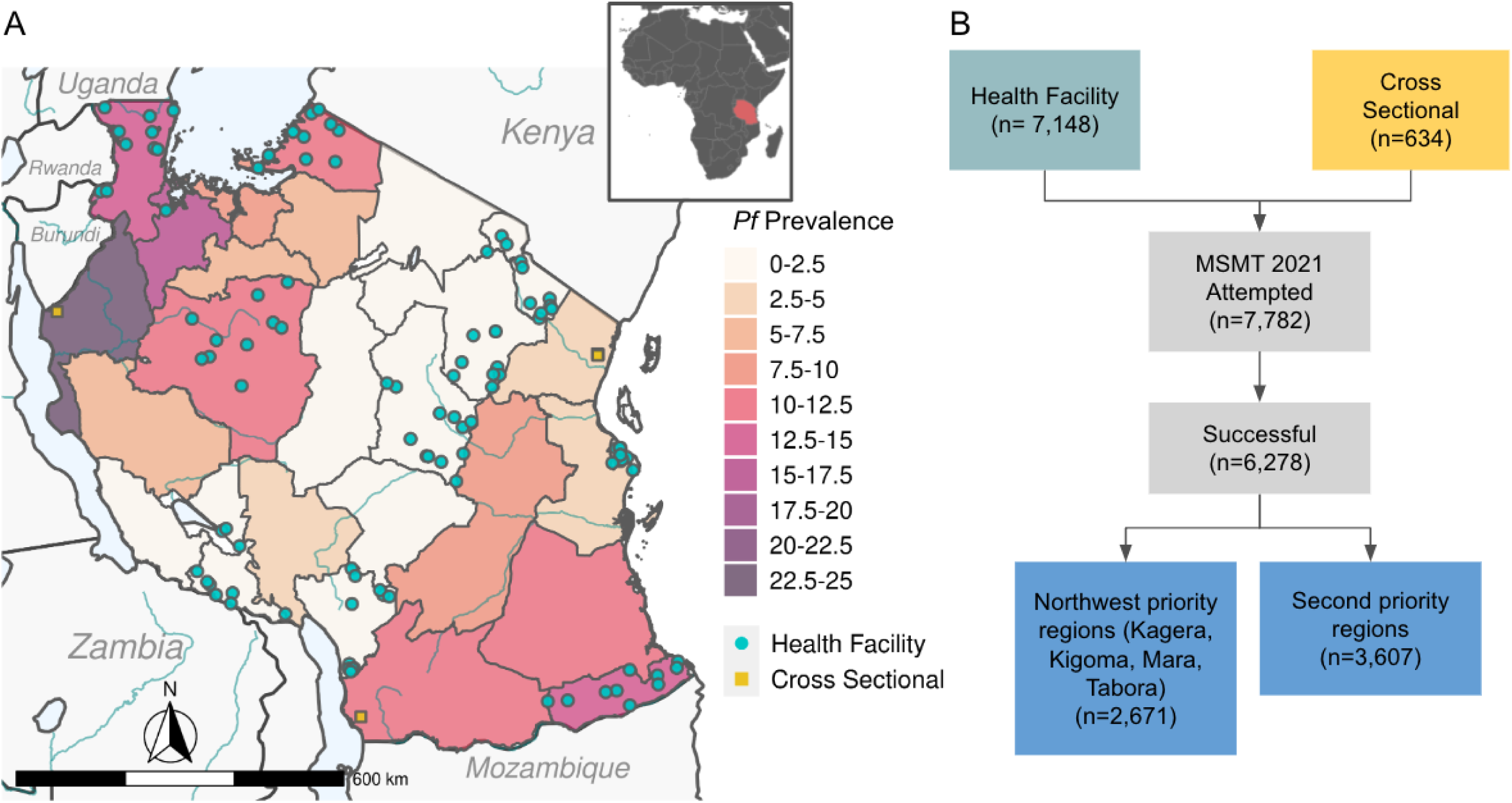
Tanzania malaria prevalence and study design. Panel A depicts a map of Tanzania showing malaria prevalence in children aged 0-59 months in 2017 as a color gradient (National Malaria Control Program data). Health facilities where sampling occurred for this study are shown as green dots, while community cross sectional collection sites are shown as yellow squares. Panel B shows the study design flowchart showing the number of samples collected at health facilities and cross sectional surveys and the success of genotyping. Priority for genotyping and analysis was given to regions in the northwest.

### Molecular Inversion Probe Analysis

DNA was extracted from the DBS using a Chelex-Tween protocol and MIP captures and sequencing were conducted as previously described.^15^ This study used a panel specific for drug resistance polymorphism detection and a panel to look at genomic diversity.^15^ Approximately two to three thousand samples were run together on each NextSeq 500 run, and sample libraries lacking sufficient read depth were rebalanced and resequenced. For samples from the high priority regions (**Figure 1**), an additional MIP capture and high-depth sequencing was conducted. Resulting data was analyzed using `MIPtools` software with `freebayes` variant calling (https://github.com/bailey-lab/MIPTools).^15^ Controls for each MIP capture and sequencing included DNA from 3D7 and 7G8 as well as no template and no probe controls.

Variant calling was conducted as previously described.^15^ We kept samples that had at least one haplotype that mapped in the expected locations of the genome for any of our drug resistance MIPs. Antimalarial resistance prevalence was calculated for all variants with a UMI count of 3 or greater and if heterozygous with the alternate allele having 1 UMI or greater using a Python script and maps were created using the sf package. Analysis of haplotypes involves only samples where complete genotypes across the involved loci are available. Inheritance by descent (IBD) analysis of parasite relatedness among ART-R parasitemias was done as previously described.^8^

### Whole Genome Sequencing

Whole genome sequencing of selective whole genome amplification (sWGA) products was attempted for 23 pure 561H, 5 mixed R561H, and 45 wildtype parasite infections based on MIP genotyping. sWGA was performed in triplicate for each sample using a previously published protocol and pooled.^19^ The pooled sWGA product was sheared using a LE220R-plus Covaris Sonicator and libraries prepared using dual indexing with the Kappa Hyper Prep Kit (Roche, Indianapolis, IN). Pooled libraries were sequenced on a NovaSeq6000 using 2 x 150 bp chemistry at the University of North Carolina (UNC) High Throughput Sequencing Facility. We also downloaded publicly available WGS data (n=25) from *P. falciparum* isolates collected in 2014/15 in Rwanda.^10^

Whole genome sequencing data was analyzed using `GATK4` following previously published methods (https://github.com/Karaniare/Optimized_GATK4_pipeline).^20^ Briefly, reads were mapped to the 3D7 reference genome using `bwa mem`, variants were called using `GATK4` and SNPs and indels were filtered using variant quality score recalibration. SNPs and indels passing filters were visualized in R 4.2.1 using the `gt` package. In order to detect patterns of selection signals between Southeast Asian (SEA), Rwanda and Tanzania haplotypes, we did extended-haplotype homozygosity (EHH) statistics focusing 561H drug resistance SNP using filtered biallelic SNPs and with low missingness data from a VCF file. All associated EHH calculations were carried out using the R-package rehh (version 2.0.4). Rwanda and SEA data was downloaded from Pf7K.

## RESULTS

We successfully genotyped 6,278 of the 7,782 (80.7%) samples attempted (**Figure 1B**). Sequencing across the K13 gene revealed three WHO validated ART-R mutations, 561H, 622I, and 675V. The 561H mutations were predominantly found in Kagera, the northwest most region bordering Rwanda and Uganda. (**Figure 2A**, **Table 1**). The overall prevalence in Kagera was 7.7% (50/649) and most of these mutant parasites were found in the west near the Rwandan border in the districts of Karagwe at 22.8% (31/136), Kyerwa at 14.4%% (17/118) and pfdhfr and pfdhps mutations were common in the area studied (Table 2). Notably, we identified Ngara, at 1.4% (2/144) (**Figure 2B**). Beyond Kagera, parasites with the 561H mutation were also detected at lower prevalence in three other regions, including Tabora at 0.5% (2/438), Manyara at 0.5% (1/179), and Njombe at 0.4% (1/279), as far as 800 km from the Rwanda border. The K13 622I mutation was found in a single isolate from Njombe in Southwestern Tanzania (**Table 1**). The 675V mutation was found in one isolate from Kagera and one isolate from Tabora. Other K13 propeller domain mutations not known to be associated with drug resistance were detected sporadically (**Table S1**). Outside the K13 propeller domain, we found several polymorphisms including 189T in 18.3% (397/2,168) of samples country-wide (**Table S1**). Other mutations have putatively been associated with partial artemisinin resistance outside of K13, including *Plasmodium falciparum* ATPase 6 (ATPase6) and *P. falciparum* AP-2mu (AP2mu) (**Table S1**). The ATPase6 623E mutation was found in four Kagera isolates and seven Mara isolates (0.6% and 2.3%, respectively), and none of these isolates contained parasites with mutations at K13 561H. *P. falciparum* coronin was not genotyped.

**Figure 2.**
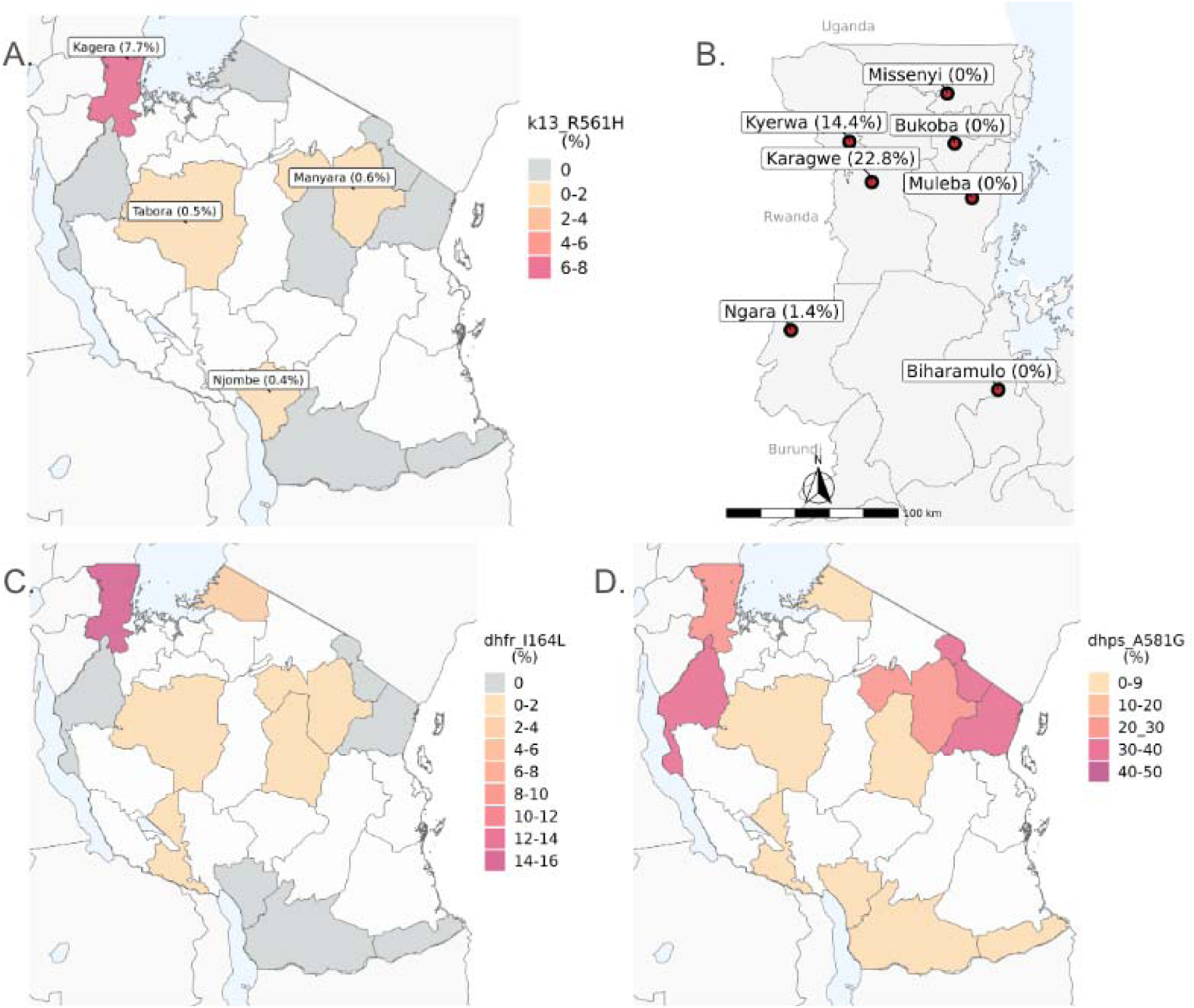
Antimalarial resistance polymorphisms in Tanzania. Panels A and B show K13 561H mutations in Tanzania by region (A) and by district in Kagera (B). In Panel A, white areas represent no sampling, gray areas represent sampled areas where no K13 561H was found, and colored areas are shaded by the frequency of samples with the 56H mutation. The red box surrounds the area plotted in Panel B, which is a map of Kagera showing K13 561H prevalence in each district sampled. DHFR 164L (Panel C) and DHPS 581G (Panel D) prevalence in sampled regions are shown in color gradients. Regions with no sampling are shown in white and regions where a polymorphism was not detected is shown in grey.

**Table 1.** Prevalence of antimalarial resistance mutations by region. Prevalence reflects the alternate allele.

| Region | DHFR<br>N51I | DHFR<br>C59R | DHFR<br>S108N | DHFR<br>I164L | DHPS<br>A437G | DHPS<br>K540E | DHPS<br>A581G | K13<br>A675V | K13<br>R561H | K13<br>R622I | MDR1<br>N86F | MDR1<br>N86Y | MDR1<br>Y184F | MDR1<br>D1246Y | CRT<br>K76T |
| --- | --- | --- | --- | --- | --- | --- | --- | --- | --- | --- | --- | --- | --- | --- | --- |
| Kagera | 0.987<br>(681/690) | 0.913<br>(630/690) | 1.0<br>(632/632) | 0.1521<br>(80/526) | 0.9408<br>(651/692) | 0.9383<br>(654/697) | 0.2593<br>(189/729) | 0.0015<br>(1/680) | 0.077<br>(50/649) | 0.0<br>(0/709) | 0.0<br>(0/223) | 0.0<br>(0/223) | 0.4936<br>(115/233) | 0.0087<br>(1/115) | 0.0164<br>(2/122) |
| Mara | 0.988<br>(574/581) | 0.926<br>(538/581) | 0.9947<br>(563/566) | 0.0305<br>(17/558) | 0.9701<br>(551/568) | 0.9724<br>(563/579) | 0.0085<br>(5/590) | 0.0<br>(0/585) | 0.0<br>(0/535) | 0.0<br>(0/578) | 0.0<br>(0/75) | 0.0<br>(0/75) | 0.337<br>(31/92) | 0.0404<br>(4/99) | 0.1395<br>(12/86) |
| Dodoma | 0.9787<br>(184/188) | 0.9149<br>(172/188) | 1.0<br>(145/145) | 0.01<br>(1/100) | 0.9509<br>(155/163) | 0.9369<br>(193/206) | 0.0543<br>(12/221) | 0.0<br>(0/182) | 0.0<br>(0/158) | 0.0<br>(0/188) | 0.0<br>(0/521) | 0.0019<br>(1/521) | 0.5204<br>(280/538) | 0.0511<br>(21/411) | 0.0052<br>(2/381) |
| Ruvuma | 1.0<br>(65/65) | 0.9846<br>(64/65) | 1.0<br>(60/60) | 0.0<br>(0/46) | 0.9444<br>(51/54) | 0.9275<br>(64/69) | 0.0278<br>(2/72) | 0.0<br>(0/68) | 0.0<br>(0/38) | 0.0<br>(0/68) | 0.0<br>(0/60) | 0.0<br>(0/60) | 0.3623<br>(25/69) | 0.0571<br>(2/35) | 0.0<br>(0/50) |
| Manyara | 0.9908<br>(216/218) | 0.9679<br>(211/218) | 1.0<br>(162/162) | 0.011<br>(1/91) | 0.9235<br>(169/183) | 0.9174<br>(200/218) | 0.2273<br>(55/242) | 0.0<br>(0/202) | 0.0056<br>(1/179) | 0.0<br>(0/202) | 0.0<br>(0/236) | 0.0<br>(0/236) | 0.5679<br>(138/243) | 0.0171<br>(2/117) | 0.0078<br>(1/128) |
| Songwe | 0.9945<br>(360/362) | 0.9613<br>(348/362) | 1.0<br>(328/328) | 0.0044<br>(1/228) | 0.9254<br>(310/335) | 0.9218<br>(342/371) | 0.0201<br>(8/399) | 0.0<br>(0/362) | 0.0<br>(0/346) | 0.0<br>(0/377) | 0.0<br>(0/377) | 0.0<br>(0/377) | 0.4197<br>(162/386) | 0.0034<br>(1/293) | 0.0<br>(0/270) |
| Tanga | 1.0<br>(58/58) | 0.9483<br>(55/58) | 1.0<br>(49/49) | 0.0<br>(0/48) | 0.9184<br>(45/49) | 0.8571<br>(54/63) | 0.3846<br>(25/65) | 0.0<br>(0/56) | 0.0<br>(0/30) | 0.0<br>(0/45) | 0.0058<br>(2/342) | 0.0058<br>(2/342) | 0.5141<br>(182/354) | 0.0036<br>(1/277) | 0.035<br>(10/286) |
| Tabora | 0.9902<br>(504/509) | 0.943<br>(480/509) | 0.9977<br>(436/437) | 0.016<br>(6/376) | 0.9725<br>(459/472) | 0.975<br>(506/519) | 0.036<br>(20/555) | 0.0022<br>(1/464) | 0.0046<br>(2/438) | 0.0<br>(0/498) | 0.0<br>(0/213) | 0.0047<br>(1/213) | 0.6197<br>(132/213) | 0.0181<br>(3/166) | 0.0331<br>(6/181) |
| Kigoma | 1.0<br>(94/94) | 0.9149<br>(86/94) | 1.0<br>(88/88) | 0.0<br>(0/88) | 1.0<br>(92/92) | 1.0<br>(87/87) | 0.3232<br>(32/99) | 0.0<br>(0/100) | 0.0<br>(0/73) | 0.0<br>(0/94) | 0.0<br>(0/215) | 0.0<br>(0/215) | 0.5411<br>(125/231) | 0.0<br>(0/70) | 0.0<br>(0/63) |
| Kilimanjaro | 0.9951<br>(202/203) | 0.9557<br>(194/203) | 1.0<br>(185/185) | 0.0<br>(0/173) | 0.9524<br>(180/189) | 0.9608<br>(196/204) | 0.3594<br>(78/217) | 0.0<br>(0/204) | 0.0<br>(0/172) | 0.0<br>(0/198) | 0.0<br>(0/556) | 0.0018<br>(1/556) | 0.637<br>(372/584) | 0.0271<br>(15/554) | 0.0054<br>(3/555) |
| Njombe | 0.991<br>(329/332) | 0.9548<br>(317/332) | 1.0<br>(291/291) | 0.0<br>(0/246) | 0.9121<br>(280/307) | 0.8991<br>(303/337) | 0.0474<br>(17/359) | 0.0<br>(0/331) | 0.0036<br>(1/279) | 0.0029<br>(1/345) | 0.0<br>(0/238) | 0.0<br>(0/238) | 0.5754<br>(145/252) | 0.0109<br>(1/92) | 0.0<br>(0/97) |
| Mtwara | 0.8602<br>(160/186) | 0.9462<br>(176/186) | 1.0<br>(107/107) | 0.0<br>(0/77) | 0.8025<br>(126/157) | 0.7857<br>(165/210) | 0.0042<br>(1/239) | 0.0<br>(0/170) | 0.0<br>(0/137) | 0.0<br>(0/144) | 0.0014<br>(1/711) | 0.0042<br>(3/711) | 0.4446<br>(325/731) | 0.0295<br>(18/610) | 0.0666<br>(39/586) |
| Dar es Salaam | 0.9548<br>(169/177) | 0.9266<br>(164/177) | 0.9914<br>(115/116) | 0.0<br>(0/49) | 0.8286<br>(116/140) | 0.8541<br>(158/185) | 0.1198<br>(26/217) | 0.0<br>(0/161) | 0.0<br>(0/137) | 0.0<br>(0/135) | 0.0<br>(0/63) | 0.0<br>(0/63) | 0.5385<br>(42/78) | 0.0<br>(0/41) | 0.0<br>(0/56) |
| Overall | 0.9817<br>(3596/3663) | 0.9378<br>(3435/3663) | 0.9984<br>(3161/3166) | 0.0407<br>(106/2606) | 0.9365<br>(3185/3401) | 0.9306<br>(3485/3745) | 0.1174<br>(470/4004) | 0.0006<br>(2/3565) | 0.017<br>(54/3171) | 0.0003<br>(1/3581) | 0.0008<br>(3/3830) | 0.0021<br>(8/3830) | 0.518<br>(2074/4004) | 0.024<br>(69/2880) | 0.0262<br>(75/2861) |

Extended haplotypes around antimalarial genes have been used to study the origin and spread of mutations.^10^ With positive directional selection of resistance by drug pressure, large genomic sections are spread in the population, a selective sweep, which are eventually broken down through recombination. We achieved sufficient depth and quality on 29 of 63 Tanzanian isolates from 2021 to assess the variation surrounding K13 and used publicly available WGS data from the isolates collected in 2014/15 in Rwanda.^10^ We identify a shared haplotype between the older Rwandan isolates and the contemporary Tanzanian isolates (n=4 from Tanzania; haplotype RW/TZ1), suggesting cross-border spread resulting from a single origin event. However, a second extended haplotype (n=5; TZ2) is also seen within the Tanzanian parasites (**Figure 3**). RW/TZ1 and TZ2 differ at the closest flanking single nucleotide polymorphisms (within 1kb of the 561H mutation). The TZ2 haplotype also does not appear to be of SEA origin (**Figure S1**). Extended haplotype homozygosity analysis (EHH) shows extended haplotypes for RW/TZ1and TZ2 relative to wild type parasites, consistent with positive selection for both 561H haplotypes (**Figure 4**). IBD analysis showed that 561H isolates from Kagera were closely related in multiple small clusters (**Figure S2**). Of the more distant isolates, the two from Tabora were related to Kagera isolates (IBD≥0.25), with the other two being more distant (**Figure S3**).

**Figure 3.**
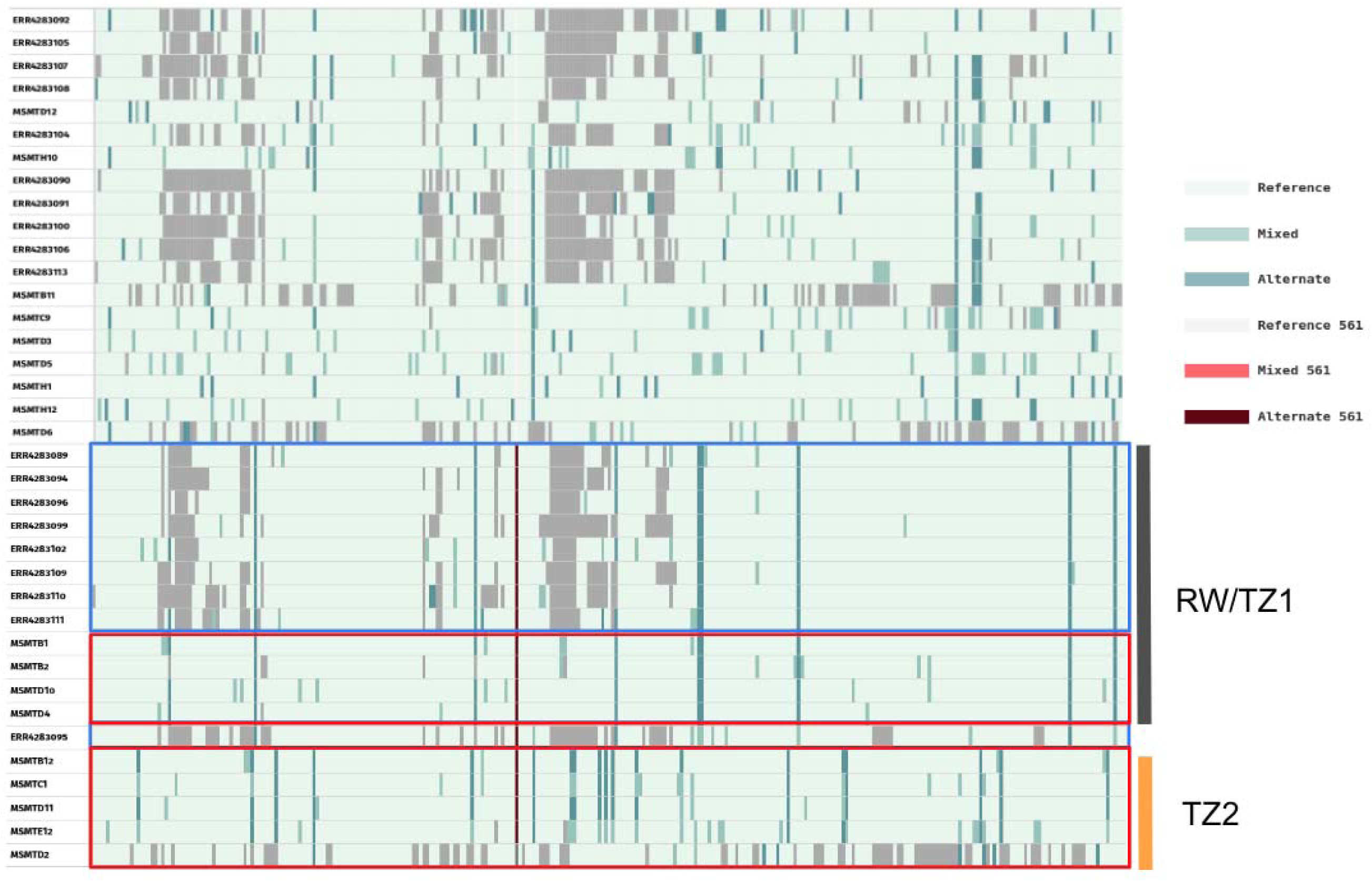
Extended flanking haplotype plot around K13 561H mutations. Pure mutant 561H are shown in maroon. Rwandan isolates are in the blue box, while Tanzanian isolates are shown in the red boxes. Grey regions represent loci where a call could not be made.

**Figure 4.**
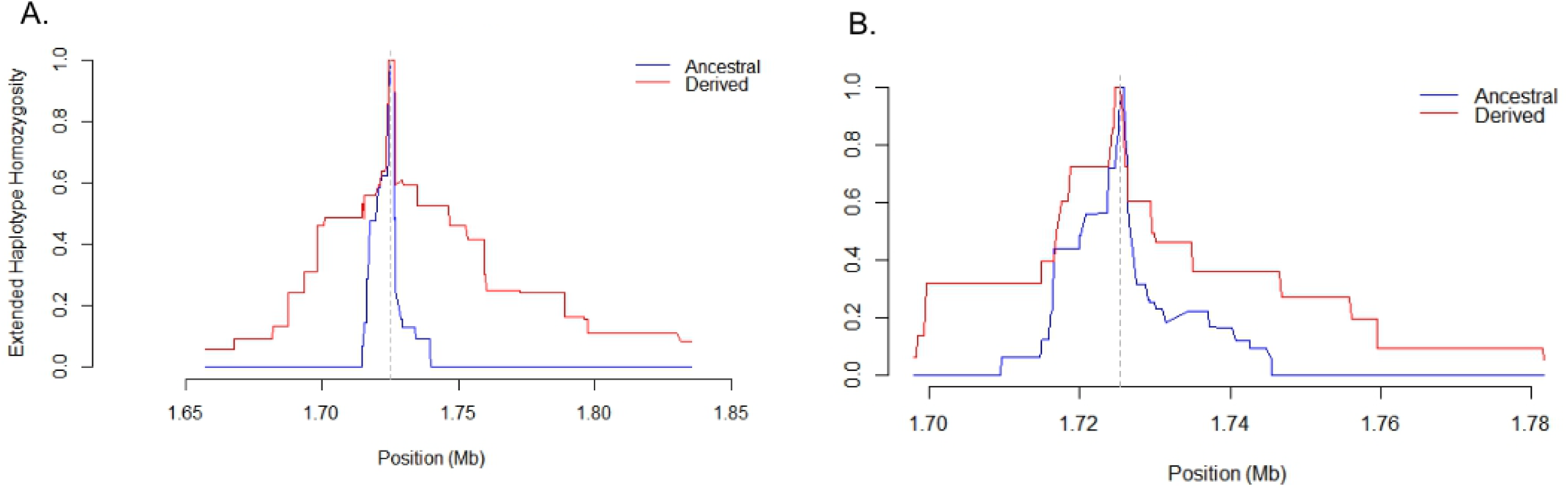
Extended Haplotype Heterozygosity (EHH) analysis of 561H haplotypes. Panel A shows the extended haplotype for the RW/TZ1 561H haplotype (red-derived) compared to wildtype parasites (blue-ancestral). Panel B shows the extended haplotype of the TZ2 561H haplotype.

To investigate associations with mutations previously found to occur in drug resistant parasites in Southeast Asia in combination with K13 mutations, we used the MIP panel genotypes and quantified mutations occurring in the same isolates. None of *P. falciparum* chloroquine resistance transporter (CRT) 326S, CRT 356T, *P.*multidrug resistance protein 2 (MDR2) 484I, *Plasmodium falciparum* Protein phosphatase (PPH) 1157L, or putative phosphoinositide-binding protein (PIB7) 1484F were associated with K13 561H mutant isolates. However, ferredoxin (FD) 193Y polymorphisms were found in 13 K13 561H mutant isolates in Kagera (**Table S1**). Known partner drug resistance mutations were also assessed (**Table 1**). In Kagera, 39 samples (39/586) had mutations at CRT 76T, and 31 of these isolates were from Ngara bordering Rwanda and Burundi (ASAQ is the most widely used ACT in Burundi). The multidrug resistance transporter 1 (MDR1) N86 allele associated with lumefantrine resistance was found in 99.9% of samples (3828/3830). The MDR1 NFD haplotype (N86, 184F, D1246) was found in 52.6% of samples (1420/2698).

Markers for SP resistance were also genotyped, revealing high frequency of many mutations and the emergence of dihydrofolate reductase (DHFR) 164L in Kagera (**Table 1**, **Figure 2C**). The folate synthesis gene triple mutations DHFR 51I, 59R and 108N were found to be near fixation with the IRN haplotype at 92.5% (2,893/3,128). Notably, the DHFR 164L mutation was found at 15.2% (80/526) in Kagera region, accounting for 80% of the IRNL haplotypes found in the country (100/2,524 nationally) (**Figure 2C**). In the dihydropteroate synthase (DHPS) gene, two markers for elevated SP resistance were found to be above WHO guideline thresholds in some regions. DHPS 540E was found at 93.06% (3,485/3,745) in Tanzania with little geographic variation (**Table 1**), while DHPS 581G prevalence ranged from 0.4% (1/239) in Mtwara to 38.5% (25/65) in Tanga (**Figure 2D**).

## DISCUSSION

Drug resistance in malaria parasites has emerged multiple times to every frontline antimalarial, each time with severe consequences for affected populations.^21^ The emerging threat of ART-R in Africa has the potential to be a global public health disaster. Thus, understanding the emergence and spread of these mutations is critical in making plans to contain this threat. While it is clear that K13 622I and 675V are spreading in the Horn of Africa and Uganda, respectively, the pattern of origin and spread of the 561H mutation have not been fully defined. Here we demonstrate that the mutation has risen to high frequencies in Kagera (7.7%), a region bordering Rwanda, which had little evidence of resistance in a 2017 study that used MIPs to genotype samples from Kagera in a national schoolkids survey (data not published). This trend would be consistent with outward spread from the Rwanda-Tanzania border, and also supports the idea that there has been and will continue to be regional spread of the mutation, especially given the shared extended haplotype around some of the Tanzanian isolates and the older reported Rwandan isolates. More concerning is the evidence of a potential unique origin of the 561H mutation in the region, suggesting that ART-R may follow patterns similar to those seen in Southeast Asia with multiple independent origins of the same mutation.^22^ Haplotype analysis suggests multiple origins, but does not prove the mechanism behind them.^6^ If multiple origins occur, containment efforts are more difficult, due to the need to closely monitor for new haplotypes and the spread of specific haplotypes, which may not become extensive without partner drug resistance. However, the presence of multiple haplotypes may be due to gene conversion or recombination events adjacent to the mutation.

Several mutations in the K13 propeller domain that are now associated with delayed parasite clearance are present in East Africa.^6–8^ The most concerning of these, 469Y, 561H, 622I and 675V, have shown clinical and *in vitro* validation of ART-R. Determining the exact origin of these mutations with certainty is difficult due to the patchy nature of the surveillance data, but one (561H) of the three appears to have originated on the Rwanda-Tanzania border. 561H was first detected in Rwanda in 2015 as part of therapeutic efficacy studies and later in DRC and Tanzania.^9,10^ To date, genotyping efforts to describe the status of K13 mutations in Tanzania have been limited and do not capture the risk posed by ART-R parasites across the country. The high rate of human movement across the border with Rwanda, where people have close historical ties and One Stop Border Posts (OSPB) that routinely permit thousands of individuals and hundreds of vehicles to cross daily, makes this a primary concern for malaria control in Tanzania.^23^ While there have been sporadic reports of validated ART-R polymorphisms in the past, systematic longitudinal surveillance of these mutations is necessary to identify areas at risk, as has been done in Uganda.^6,9,17^ The speed at which data is generated and reported to control programs is critical. The MSMT project will provide these nationwide data for Tanzania given the identified threat shown here of 561H in Kagera, by allowing yearly sampling and in-country sequencing and analysis of data as the project develops.

Partner drug resistance surveillance showed important patterns. An intriguing pattern of CRT 76T mutations was found to be clustered near the Burundi border. A possible explanation is that the use of ASAQ in Burundi is contributing to continued selection for chloroquine resistance and that these resistant parasites are being imported into Kagera. Given artemether-lumefantrine remains the first line antimalarial in Tanzania, it is not surprising that MDR1 N86 was near fixation and the NFD haplotype was very common, occurring in over 50% of samples.

Markers of elevated SP resistance are increasing in Tanzania, which has potential impacts on the use of SP as a chemopreventive antimalarial. The geographic distribution of the DHPS 581G mutations in Tanzania has historically been limited to Tanga and the Lake districts.^24,25^ We confirm that this remains the case with little evidence of expansion into other regions of Tanzania. The reasons for the lack of spread of this mutation in Tanzania remains unclear and warrants further evaluation, especially given the evidence that spread is occurring in the neighboring Democratic Republic of the Congo.^26^ Previous work has suggested that multiple origins of parasites bearing 581G have occurred in East Africa, suggesting local origins and limited spread.^27^ However, that work was based on five microsatellites and may not take into account the complex stepwise evolutionary history of mutations in DHPS. Importantly, a new focus of the quadruple mutation in DHFR (51I, 59R, 108N, 164L) was identified in Kagera. In the laboratory, this form of the enzyme binds pyrimethamine 600 times less tightly than the wild type and about seven times less than the triple mutation (51I, 59R, 108N).^28^ This results in parasites being resistant to therapy *in vitro* at levels higher than what can be reached *in vivo*. The DHFR/DHPS sextuple mutation (DHFR 51I, 59R, 108N / DHPS 437G, 540E, 581G), has been associated with the compromise of intermittent preventive treatment in pregnancy (IPTp) and was found in 11.2% (323/2,894) of samples nationally.^29^ Given the regional high prevalence of DHPS 581G and the emergence of DHFR 164L in Northwest Tanzania, additional studies are warranted to evaluate the benefits of SP based chemoprevention in this region of Tanzania.

A major advantage of this study is the large geographic range and large number of samples available for analysis. However, this study design also engenders some limitations. The large number of samples resulted in overall low levels of coverage for the sequencing for most regions (other than the four high priority regions that received extra depth). The samples collected at the three cross-sectional sites performed poorly with our approach, likely due to low parasitemia. The overall coverage in many regions may directly affect the ability to find minority drug resistant variants in the data and we may have filtered out minor variants below 1%.

The detection of K13 561H at prevalence above 22% in Karagwe district of Kagera, Tanzania, must raise alarms thatART-R is emerging and that ACTs efficacy is under threat. The independent origin of a new K13 561H extended haplotype raises concern that multiple origins of ART-R mutations will occur in Africa, complicating control. Evolution of partner drug resistance must be monitored carefully and therapeutic efficacy studies are urgently needed to understand the susceptibility of currently circulating K13 mutations in the region.

## Authors contribution

DSI, JJJ and JB formulated the original idea. DSI, CIM, RAM, MDS and CB conducted the field surveys. DG, AF, AS, BL, ZPH, KN and AL performed data analysis under the guidance of DSI, JJJ and JB. JJJ, AS, AF, DG, BL, ZPH, CB, DP, MDS, JB, and DSI wrote and edited the manuscript. All authors contributed to the article and approved the submitted version.

## Acknowledgements

The authors wish to thank participants and parents/guardians of all children who took part in the surveillance. We acknowledge the contribution of the following project staff and other colleagues who participated in data collection and/or laboratory processing of samples; Raymond Kitengeso, Ezekiel Malecela, Muhidin Kassim, Athanas Mhina, August Nyaki, Juma Tupa, Anangisye Malabeja, Emmanuel Kessy, George Gesase, Tumaini Kamna, Grace Kanyankole, Oswald Osca, Richard Makono, Ildephonce Mathias, Godbless Msaki, Rashid Mtumba, Gasper Lugela, Gineson Nkya, Daniel Chale, Richard Malisa, Sawaya Msangi, Ally Idrisa, Francis Chambo, Kusa Mchaina, Raymond Kitengeso, Neema Barua, Christian Msokame, Rogers Msangi, Salome Simba, Hatibu Athumani, Mwanaidi Mtui, Rehema Mtibusa, Jumaa Akida, Ambele Yatinga, Tilaus Gustav, Oksana Kharabora and Claudia Gaither. The finance, administrative and logistic support team at NIMR: Christopher Masaka, Millen Meena, Beatrice Mwampeta, Gracia Sanga, Neema Manumbu, Halfan Mwanga, Arison Ekoni, Twalipo Mponzi, Pendael Nasary, Denis Byakuzana, Alfred Sezary, Emmanuel Mnzava, John Samwel, Daud Mjema, Seth Nguhu, Thomas Semdoe, Sadiki Yusuph, Alex Mwakibinga, Rodrick Ulomi and Andrea Kimboi. Management of the National Institute for Medical Research, National Malaria Control Program and President’s Office-Regional Administration and Local Government (Regional administrative secretaries of the 13 regions, and district officials, staff from all 100 HFs and Community Health Workers from the 3 community cross sectional regions. Technical and logistics support from the Bill and Melinda Gates Foundation team is highly appreciated. Permission to publish the manuscript was sought and obtained from the Director General of NIMR.

## Data Availability

All sequencing data has been submitted to SRA (# pending). Other metadata is available from the corresponding author upon reasonable request. Code for analysis is available at: https://github.com/bailey-lab/MSMT_2021_DR_analyses.

