## Supplemental Material for "Country wide surveillance reveals prevalent artemisinin partial resistance mutations with evidence for multiple origins and expansion of high level sulfadoxine-pyrimethamine resistance mutations in northwest Tanzania"

**Supplementary Table 1. Mutations in antimalarial resistance genes**.

See uploaded Excel file.

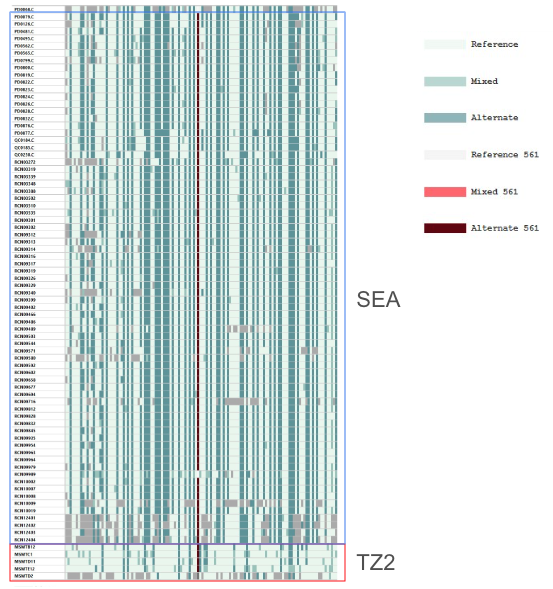

**Figure S1. Extended haplotype comparison between TZ2 and Southeast Asian (SEA) isolates.** 561H mutant parasites collected in SEA from Pf7K (blue box) are compared to TZ2 isolates (red box) from this study. Extended haplotypes show two distinct groups.

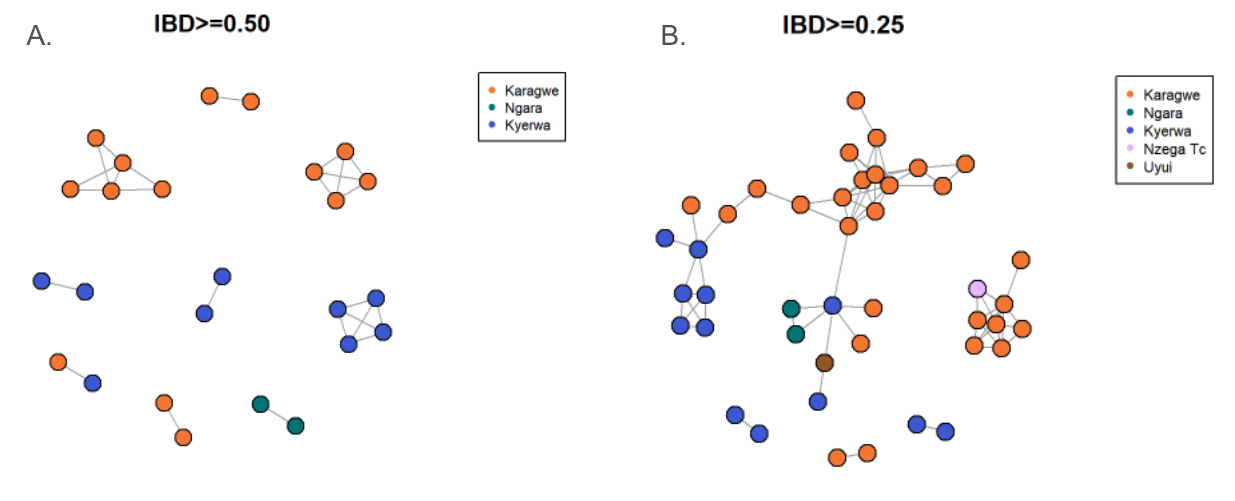

**Figure S2. Inheritance by descent relatedness among 561H parasite infection in Kagera.** IBD ranges from 0 (completely unrelated) to 1 (identical) for infections. Panel A and B show networks of parasites with 0.5 and 0.25 IBD, respectively. Isolates are colored by the district that they are from.

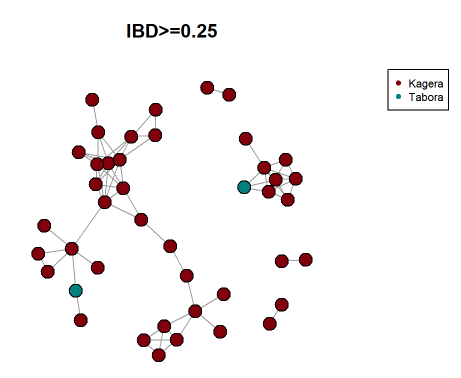

**Figure S3. IBD relatedness of 561H infections between regions.** At an IBD level of 0.25, the two parasite isolates from Tabora cluster with isolates from Kagera. The isolates in other regions remain not connected to the network demonstrating lower levels of relatedness.
